# Vascular Risk Factor Prevalence and Trends in Native Americans With Ischemic Stroke - A National Inpatient Sample Analysis

**DOI:** 10.1101/2020.10.16.20214148

**Authors:** Dinesh V. Jillella, Sara Crawford, Rocio Lopez, Atif Zafar, Anne S. Tang, Ken Uchino

## Abstract

**Introduction:** Native Americans have a higher incidence and prevalence of stroke and the highest stroke-related mortality among race-ethnic groups in the United States. We aimed to analyze trends in the prevalence of vascular risk factors among Native Americans with ischemic stroke over the last two decades along with a comparison to the other race-ethnic groups.

**Methods:** National/Nationwide Inpatient Sample (NIS) database was used to explore the prevalence of risk factors among hospitalized ischemic stroke patients during 2000 - 2016. Ischemic stroke and risk factors of interest were identified using validated ICD-9/10 codes. The race-ethnic groups of interest were Native American, White, Black, Hispanic, Asian/Pacific Islanders, and others. Crude and age-and sex-standardized prevalence estimates were calculated for each risk factor within each race-ethnic group in 6 time periods: 2000-02, 2003-05, 2006-08, 2009-11, 2012-14, and 2015-16. We explored linear trends over the defined time periods using linear regression models, with differences in trends between the Native American group and each of the other race-ethnic groups assessed using interaction terms. The analysis accounted for the complex sampling design, including hospital clusters, NIS stratum, and trend weights for analyzing multiple years of NIS data.

**Results:** Of the 1,278,784 ischemic stroke patients that were included in the analysis, Native Americans constituted 5472. The age-and-sex-standardized prevalence of hypertension (trend slope = 2.24, p < 0.001), hyperlipidemia (trend slope = 6.29, p < 0.001), diabetes (trend slope = 2.04, p = 0.005), atrial fibrillation/flutter (trend slope = 0.80, p = 0.011), heart failure (trend slope = 0.73, p = 0.036) smoking (trend slope = 3.65, p < 0.001), and alcohol (slope = 0.60, p = 0.019) increased during these time periods among Native Americans, while coronary artery disease prevalence remained unchanged. Similar upward trends of several risk factors were noted across other race-ethnic groups with Native Americans showing larger increases in hypertension prevalence compared to Blacks, Hispanics, and Asian/Pacific Islanders and in smoking prevalence compared to Hispanics and Asian/Pacific Islanders. By the year 2015-2016, Native Americans had the highest overall prevalence of diabetes, coronary artery disease, smoking, and alcohol among all the race-ethnic groups.

**Conclusion:** The prevalence of most vascular risk factors among ischemic stroke patients has increased in Native Americans and other race-ethnic groups over the last two decades. Significantly larger increases in the prevalence of hypertension and smoking were seen in Native Americans compared to other groups along with them having the highest prevalence in multiple risk factors in recent years.

## Introduction

Although global mortality rates for stroke have improved over the last three decades, stroke continues to be the second leading cause of death worldwide^1,2^. An update from the Global Burden of Disease Study shows the relative increase in lifetime stroke risk from 1990 to 2016 to be about 8.9% ^3^. Racial-ethnic variability has also remained a concern with some racial-ethnic groups experiencing excess disease burden compared to the others^4^. Native Americans, in particular, are reported to have a higher incidence and prevalence of stroke than Whites and Blacks and the highest stroke-related mortality out of all racial and ethnic groups in the United States ^5-7^. Approximately 90% of the cerebrovascular risk can be attributed to modifiable risk factors like hypertension, hyperlipidemia, and diabetes ^4^. While it is reported that Native Americans are disproportionately affected by several cerebrovascular risk factors compared to their racial-ethnic counterparts based on some smaller studies ^5,8,9^, there is a paucity of data detailing recent trends in burden.

We aimed to analyze trends in the prevalence of vascular risk factors among Native Americans with ischemic stroke (IS) over the last two decades and compare it with the other race-ethnic groups during the same period.

## Methods

We used the National Inpatient Sample and the Nationwide Inpatient Sample (NIS) database from the Healthcare Cost and Utilization Project (HCUP), Agency for Healthcare Research and Quality (AHRQ) for 2000-2016 ^10,11^. The prevalence of risk factors by race-ethnicity was explored during this period among adult inpatient hospital discharges for ischemic stroke, identified using ICD-09 codes of 434.01, 434.11, 434.91, 433.01, 433.11, 433.21, 433.31, 433.81, 433.91, and 436 in addition to ICD-10 codes of I63^12^. Risk factors of interest were also identified using validated ICD-09 and ICD-10 codes, included essential hypertension, diabetes mellitus, hyperlipidemia, atrial fibrillation or flutter, heart failure, coronary artery disease, alcohol use, and smoking/tobacco abuse. The race-ethnic categories of interest included non-Hispanic White, non-Hispanic Black, Hispanic, Asian/Pacific Islander, Native American, and other. We excluded individuals whose race-ethnic information was missing.

This study was approved under a waiver of informed consent by the Cleveland Clinic institutional review board.

### Statistical analysis

We calculated both crude and age- and sex-standardized prevalence estimates for each risk factor across all years by race-ethnicity. We then stratified the results by time period, exploring the periods 2000-02, 2003-05, 2006-08, 2009-11, 2012-14, and 2015-16. Direct age and sex standardization was performed using 2000 census data as the standard population ^13^. We report weighted prevalence estimates and 95% confidence intervals. Linear trends over the defined time periods were assessed using linear regression models, with differences in trends between the Native American group and each of the other race-ethnic groups assessed using interaction terms. Estimates derived from an unweighted frequency of less than 10 or with a coefficient of variation of more than 0.30 are suppressed due to lack of stability. The analysis accounted for the complex sampling design, including hospital clusters, NIS stratum, and trend weights for analyzing multiple years of NIS data^14-16^. All tests were two-tailed and performed at a significance level of 0.05 using SAS version 9.4 (The SAS Institute, Cary, NC).

## Results

A total of 1,534,300 adult ischemic stroke patient discharges were captured during the years 2000-2016 from the NIS database, representing 7,406,6380 weighted hospitalizations. Of these, race-ethnicity information was available in 1,278,784 (83.3%) representing 6,183,757 weighted hospitalizations. Native Americans constituted 5,472 hospitalizations (weighted frequency (percent) of 26,775 (0.44%)) of these with females constituting 2,856 (weighted frequency (percent) of 13,986 (52.25%)) of the Native Americans (Table 1).

**Table 1:** Characteristics of overall racial distribution and age and gender prevalence of Native American Ischemic Stroke Patients, 2000-2016.

| Category | N Unweighted | N Weighted | % Weighted |
| --- | --- | --- | --- |
| **Total** | 1,278,784 | 6,183,757 |  |
| **Race** |  |  |  |
| Native American | 5,472 | 26,775 | 0.43 |
| White | 909,494 | 4,399,398 | 71.15 |
| Black | 205,903 | 996,087 | 16.11 |
| Hispanic | 93,753 | 450,929 | 7.29 |
| Asian/Pacific Islander | 33,375 | 161,218 | 2.61 |
| Other | 30,787 | 149,350 | 2.42 |
| **Age distribution among Native Americans** |  |  |  |
| < 45 years | 326 | 1,597 | 5.96 |
| 45 to 54 years | 740 | 3,621 | 13.52 |
| 55 to 64 years | 1,232 | 6,008 | 22.44 |
| 65 to 74 years | 1,337 | 6,542 | 24.43 |
| 75 to 84 years | 1,216 | 5,964 | 22.27 |
| ≥85 years | 621 | 3,043 | 11.37 |
| **Gender distribution among Native Americans** |  |  |  |
| Male | 2,614 | 12,780 | 47.75 |
| Female | 2,856 | 13,986 | 52.25 |

From 2000-2016, the crude prevalence of vascular risk factors among Native Americans increased with respect to hypertension (slope = 1.51, p = 0.003), hyperlipidemia (trend slope = 7.54, p < 0.001), diabetes (trend slope = 1.93, p = 0.005), smoking (trend slope = 3.04, p < 0.001), atrial fibrillation/flutter (trend slope = 0.87, p = 0.011), and alcohol (trend slope = 0.51, p = 0.022) during these time periods, while coronary artery disease and heart failure prevalence remained unchanged.

The age- and sex-standardized prevalence of hypertension significantly increased from 47.96% to 64.32 % and diabetes from 44.16% to 51.79% among Native American patients during 2000-2016 stratified by 6 time periods (Table 2). During this time, hyperlipidemia, atrial fibrillation, and heart failure more than doubled in their prevalence with estimates increasing from 23.71% to 53.22%, 3.77% to 8.67%, and 4.42% to 11.50%, respectively. Smoking prevalence almost doubled, showing an increase from 21.62% to 40.69%. Although coronary artery disease showed a numerical increase from 10.56% to 16.37% during this period, the trend was not statistically significant, as prevalence fluctuated over time.

**Table 2:** Age-and-sex-standardized vascular risk factor prevalence in Native Americans from 2000 to 2016.

| **Risk Factor** | **2000-02** | **2003-05** | **2006-08** | **2009-11** | **2012 - 14** | **2015 - 16** | **Trend Slope** | **p-value** |
| --- | --- | --- | --- | --- | --- | --- | --- | --- |
| Hypertension | 47.96 | 54.32 | 59.96 | 59.33 | 63.47 | 64.32 | 2.24 | <0.001 |
| Hyperlipidemia | 23.71 | 33.59 | 40.48 | 35.53 | 52.00 | 53.22 | 6.29 | < 0.001 |
| Diabetes mellitus | 44.16 | 51.87 | 44.71 | 40.33 | 45.39 | 51.79 | 2.04 | 0.005 |
| Coronary artery disease | 10.56 | 14.91 | 18.34 | 15.36 | 14.56 | 16.37 | 0.49 | 0.23 |
| Atrial fibrillation or flutter | 3.77 | 5.02 | 5.75 | 6.32 | 6.99 | 8.67 | 0.80 | 0.01 |
| Heart Failure | 4.42 | 10.48 | 7.35 | 9.22 | 9.45 | 11.50 | 0.73 | 0.03 |
| Smoking | 21.62 | 17.84 | 27.62 | 30.93 | 27.80 | 40.69 | 3.65 | <0.001 |
| Alcohol | 5.16 | 8.73 | 8.22 | 8.75 | 8.12 | 8.61 | 0.60 | 0.019 |

The age- and sex- standardized prevalence increased in most vascular risk factors in all the race-ethnic groups (Figure). Hypertension showed a significant increase in prevalence among Whites (Slope=1.39, p= <0.001), Blacks (Slope=0.57, p<0.001), and Hispanics (0.80, p<0.001) (Figure Panel A). No significant upward trend was found in Asian/Pacific Islanders (Slope=0.23, p=0.38). Native Americans experienced a significantly steeper trend (slope=2.24, p<.0001) in comparison to Blacks, Hispanics, and Asian/Pacific Islanders (difference in trends of 1.67 (p=0.003), 1.44 (0.011), and 2.01 (p=0.0009), respectively).

Hyperlipidemia showed a significant increase in the adjusted prevalence in all the race-ethnic groups (Figure Panel B). Native Americans had a slope for a trend of 6.29, Whites 5.71, Blacks 6.24, Hispanics 6.53, and Asian/Pacific Islander 5.59, with p<0.0001 for all. The trend among Native Americans was not significantly different than the other race-ethnic groups although they had the highest net difference in hyperlipidemia prevalence after Hispanics. Similar to hyperlipidemia, diabetes mellitus also showed a significant increase in adjusted prevalence in all the race-ethnic groups (Figure Panel C). Native Americans had the steepest upward trend during the 16 years, but the trend was not significantly different than the trends in the other race-ethnic groups. Coronary artery disease (Figure Panel D) was the only vascular risk factor that did not show an increase in the prevalence over time among Native Americans, but Whites and Blacks did show an increase with slopes of 0.23 (p<0.0001) and 0.81 (p<0.0001), respectively.

The adjusted prevalence of atrial fibrillation/flutter (Figure Panel E) more than doubled in Native Americans during the 16-year time period from 3.77% to 8.67% (slope=0.80, p=.011). Increasing trends were also identified across all the other race-ethnic groups of interest during this period. The difference in trends across Native Americans and other races was non-significant. The adjusted prevalence of heart failure in Native Americans also more than doubled, (Figure Panel F) increasing from 4.42% to 11.50% (slope=0.73, p=0.036). Significant upward trends were noted across all the race-ethnic groups, and interestingly, the Black population had a significantly steeper slope at 1.47 (p<.001) compared to Native Americans (difference in slopes=0.74, p=0.038).

Native Americans showed a significant uptrend in both alcohol use and smoking, with increases from 5.16% to 8.61% (slope=0.60, p=0.019) and 21.62% to 40.69% (slope=3.65, p<.001), respectively (Figure Panels G and H). The adjusted prevalence among Native Americans was the highest of all the race-ethnic groups in these two categories by the end of the study period in 2016; however, their upward trend was only significantly steeper than Hispanics and Asian/Pacific Islanders. By the year 2015-16, Native Americans had the highest adjusted prevalence of diabetes (64.3%), coronary artery disease (16.37%), smoking (40.69%), and alcohol use (8.61%) across all race-ethnic groups and were in the highest tertiles with regards to hyperlipidemia, heart failure, and hypertension.

## Discussion

In our study, the age- and sex-standardized prevalence of all ischemic stroke risk factors increased significantly in Native Americans during 2000-2016 except for coronary artery disease. Increases in prevalence were also seen in the other race-ethnic groups, but the Native Americans showed larger uptrends in hypertension compared to Blacks, Hispanics, and Asian/Pacific Islanders and in smoking compared to Hispanics and Asian/Pacific Islanders. Of note, Native Americans showed the highest prevalence percentages in multiple vascular risk factors in recent years compared to their counterparts.

Native Americans in the US constitute a very diverse group and have many reasons for health disparity. A few include geographic isolation, limitation of transportation, economic disparities, and language, educational, and cultural barriers^17^. Such factors could potentially influence the prevalence of cerebrovascular and cardiovascular diseases. Data from the mid-1900s suggested lower cardiovascular and cerebrovascular disease prevalence and mortality among Native Americans^18^. However, with improving race-ethnicity identification measures and classifications starting in the early 1990s, the cardiovascular and cerebrovascular disease prevalence and mortality were found to be higher in the Native American populations^7,19^. The reported prevalence of stroke in Native Americans is almost twice that of White Americans, with ischemic stroke constituting >85% of these^5,6^. In this setting, the identification and management of vascular risk factors that are potentially contributory to this along with the utilization of preventative strategies are of utmost importance. Our study showed concerning increases in the risk factors in the Native American population over the last two decades. Of additional concern, is the observation of the highest prevalence of multiple stroke vascular risk factors in recent years, especially modifiable ones like diabetes, smoking, and alcohol.

Substance abuse continues to be an area of concern within the Native American population. The almost doubling of smoking percentages over a 16-year time period to an adjusted prevalence of more than 40% among the Native American ischemic stroke patient population in our study is striking. This is in contrast to a concurrent decline in smoking rates within the general U.S. adult population, which fell from 20.9% in 2005 to 15.5 % in 2016, according to data from the United States Centers for Disease Control and Prevention^20^. Although not as drastic as smoking, alcohol also continues to increase in the Native American population, with the adjusted prevalence greater than that of all other race-ethnic groups by the end of the study period.

Of note, although increasing trends in the vascular risk factors were noted universally across the study period, minorities, especially Asian/Pacific Islanders and Blacks, had the highest prevalence of vascular risk factors by the end of the study period with Asian/Pacific Islanders having the highest prevalence in hypertension, hyperlipidemia, and atrial fibrillation/flutter closely followed by the Native Americans in each of these categories. Blacks had the highest prevalence of heart failure by the end of the study period in 2016.

A recent study by Otite et al. reported increases in the prevalence of hypertension, diabetes, dyslipidemia, smoking, and drug abuse in acute ischemic stroke patients across a 10-year time period^21^. Our study confirmed this trend over 16 years, and when stratified across multiple race-ethnic groups, showed that the Native Americans as a population have done relatively worse compared to their counterparts. Identification of such increases in vascular risk factors, especially in specific race-ethnic and minority groups with health and socioeconomic disparities, can help us target these risk factors as a means of reducing stroke burden going forward. Aggressively modifying these risk factors at the grassroots level can help in primary and secondary stroke prevention, especially in underprivileged communities with limited access to health resources like the Native Americans, Blacks, and Asian/Pacific Islanders.

Most of the information on Native Americans especially pertaining to their vascular risk factors is based on the “The Strong Heart Study” data^5,22^. Based on this large prospective study utilizing data up to early 2000s, incidence and case-fatality rates of stroke in Native Americans were high compared to others and strong associations between hypertension, diabetes, and cigarette smoking and risk of stroke were found^5^. Of note, earlier data from this study over a 4-year period between 1989 to 1995 showed a decrease in smoking and alcohol use rates, at the same time showing an uptrend in hypertension and diabetes prevalence. In this setting, our study showing recent uptrends in all the vascular risk factors is of vital importance.

The inclusion of a large sample of more than 5000 Native American ischemic stroke patients is a major strength of our study especially considering the dearth of recent data on vascular risk factors in this race-ethnic group. Considering its retrospective analysis of administrative data, there could be bias related to coding errors and misclassifications. A combination of ICD-9 and recent ICD-10 diagnostic codes was used in this study. Secular trends in the documentation and coding of comorbidities due to electronic health record use or to increase reimbursement could have affected the prevalence of vascular risk factors although with variable data^23,24^. Considering the limitation of the NIS database in tracking re-admissions, recurrent admissions of a single individual, could result in duplication of entries. In the same way, interhospital transfers after hospitalization may also lead to duplicate registrations of a patient ^25^. The category of Native Americans (and Alaskan Natives) is generalized and the data are not stratified by different tribes or by their geographical location. Also, the race-ethnic categorization across health care facilities in the U.S. over time has not been validated. Lastly, race-ethnicity information was missing in about 17% of the data leading to their exclusion from our study, which is another limitation.

## Conclusion

The prevalence of most vascular risk factors among ischemic stroke patients has increased in Native Americans and other race-ethnic groups over the last two decades. Native Americans have experienced significantly larger increases in hypertension and smoking and have the highest prevalence in multiple risk factors in recent years, as compared to other race-ethnic groups. These results highlight the need for stroke prevention by aggressive stroke risk factor management and by targeting individual cerebrovascular risk factors in minorities like the Native Americans, Blacks, and Asian/Pacific Islanders. Further large-scale prospective studies are necessary to better define interventions, which can help improve the health care of minority populations who are limited in their access to quality health care resources.

## Data Availability

NIS data are de-identified and publicly available through AHRQ HCUP (www.hcup-us.ahrq.gov)

## Acknowledgments

This study was possible with the support of the Center for Population Health Research, Cleveland Clinic Foundation.

---

**FIGURE:**
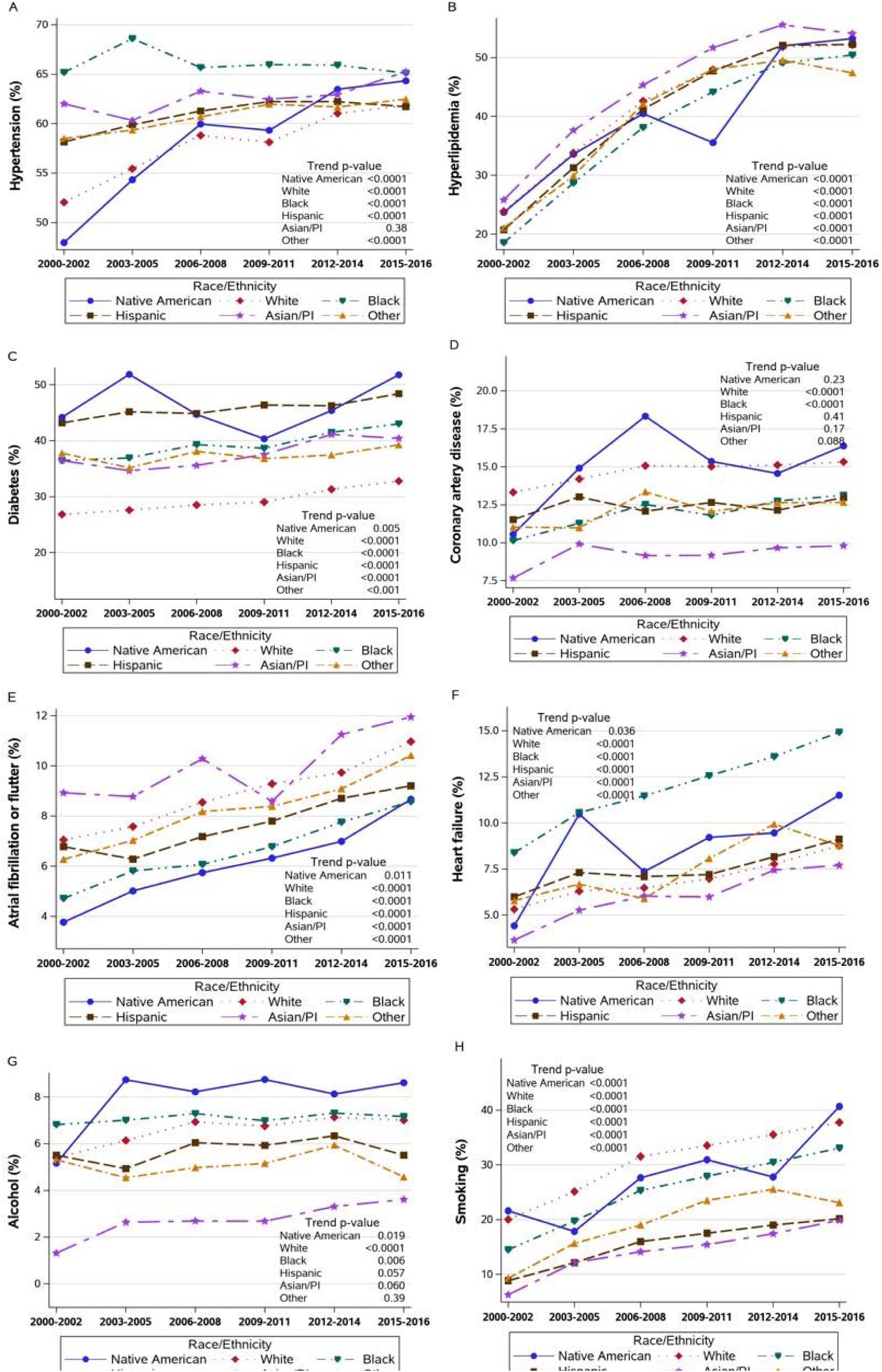
Age-and-Sex-standardized vascular risk factor prevalence in Native Americans and other race-ethnic groups from 2000 to 2016. A: Hypertension prevalence across race-ethnic groups B: Hyperlipidemia prevalence across race-ethnic groups C: Diabetes prevalence across race-ethnic groups D: Coronary artery disease prevalence across race-ethnic groups E: Atrial fibrillation or flutter prevalence across race-ethnic groups F: Heart failure prevalence across race-ethnic groups G: Alcohol prevalence across race-ethnic groups H: Smoking prevalence across race-ethnic groups

